# CD8 and vimentin were associated with overall survival in patients with ovarian cancer treated with intraperitoneal chemotherapy

**DOI:** 10.1101/2024.12.27.24319705

**Authors:** Hyojin Kim, Ji Eun Lee, Yong Jae Lee, Kidong Kim

## Abstract

**Objective:** To identify immunohistochemistry markers affecting the survival in patients with ovarian cancer receiving intraperitoneal (IP) chemotherapy.

**Methods:** A retrospective review of medical records identified 24 patients with newly diagnosed stage III or IV high-grade serous ovarian carcinoma who underwent more than three cycles of IP chemotherapy at a tertiary hospital in Republic of Korea between 1990 and 2013. Immunohistochemical staining of tumor tissue for CD8, FOXP3, PDL1, E-cad and vimentin was performed. The level of expression was measured using established protocols of each marker and was dichotomized (high vs. low) using median value. The association of level of expression of each marker with progression-free survival (PFS) or overall survival (OS) were examined.

**Results:** The mean age was 44 years (range 27 to 59) and 23 patients were stage III. The median PFS was 15.3 months (range 0.4 to 148.3) and that of OS was 63.3 months (range 0.4 to 163.0). None of 5 markers were associated with PFS. However, CD8 (p=0.2) and vimentin (p=0.1) were marginally associated with OS. Patients with high CD8 or vimentin expression demonstrated a numerically longer PFS compared to those with low expression of both markers (median 19.7 months vs. 13.0 months, *p* = 0.073). Furthermore, patients with high CD8 or vimentin expression showed significantly improved OS compared to those with low expression of both markers (median 94.5 months vs. 25.4 months, *p* = 0.008).

**Conclusion:** CD8 and vimentin expression were correlated with OS in patients with ovarian carcinoma treated with IP chemotherapy.

## Introduction

Ovarian cancer is a highly aggressive gynecologic malignancy with increasing incidence and mortality rates in Korea [1]. Despite advancements in surgical techniques and chemotherapeutic agents, the majority of patient experience disease recurrence within 12 to 18 months. Among patients with platinum-refractory disease, median survival does not exceed 5 months [2,3].

The peritoneal cavity is the primary site of metastasis and recurrence in epithelial ovarian cancer, rendering intraperitoneal (IP) chemotherapy a targeted approach for drug delivery. Studies have reported that IP chemotherapy confers a survival benefit compared to intravenous (IV) chemotherapy [4] and the NCCN Guidelines recommend the IP/IV regimen as a treatment option for ovarian cancer [5,6]. However, subsequent trials failed to demonstrate the superiority of IP chemotherapy over IV chemotherapy in this patient population [4]. Furthermore, IP chemotherapy is associated with adverse effects such as abdominal pain, dermatitis, peritonitis, and catheter-related complications. Consequently, the role of IP chemotherapy in the management of ovarian cancer remains a subject of ongoing debate. We previously reported a case in which a patient with recurrent epithelial ovarian cancer, resistant to multiple lines of intravenous chemotherapy, achieved complete remission with IP chemotherapy [7]. This observation led us to hypothesize that IP chemotherapy may be effective in a specific subset of ovarian cancer patients. To identify this subset, we investigated the association between immunohistochemical markers and survival outcomes in patients with ovarian cancer who received adjuvant IP chemotherapy.

## Materials and Methods

### Study population

This retrospective study utilized data from a tertiary hospital in Republic of Korea collected between January 1, 1990, and January 31, 2013. The study focused on women aged 19 years and older with a newly diagnosed primary epithelial ovarian, fallopian tube, or peritoneal cancer who underwent primary cytoreductive surgery followed by IP chemotherapy. Routine surveillance included serial CA-125 measurements and imaging studies. The institutional follow-up protocol involved patient evaluations every 3 months during the first 2 years post-treatment and every 6 months thereafter. Recurrence was defined as the date of radiologically confirmed disease detected during follow-up. An isolated elevation of CA-125 in the absence of clinical or radiological evidence of relapse was not classified as progression but prompted further radiological assessments. From an initial cohort of 35 patients, this study focused on those with high-grade serous ovarian carcinoma, excluding patients diagnosed with endometrioid carcinoma (n = 2) and clear cell carcinoma (n = 4). Patients included in the final analysis had FIGO stage III or IV disease and received more than four cycles of IP chemotherapy following primary surgery, resulting in a final study population of 24 patients (Fig. 1). This study was approved by the Institutional Review Board of our institution (IRB NO B-1912-582-305).

**Figure 1.**
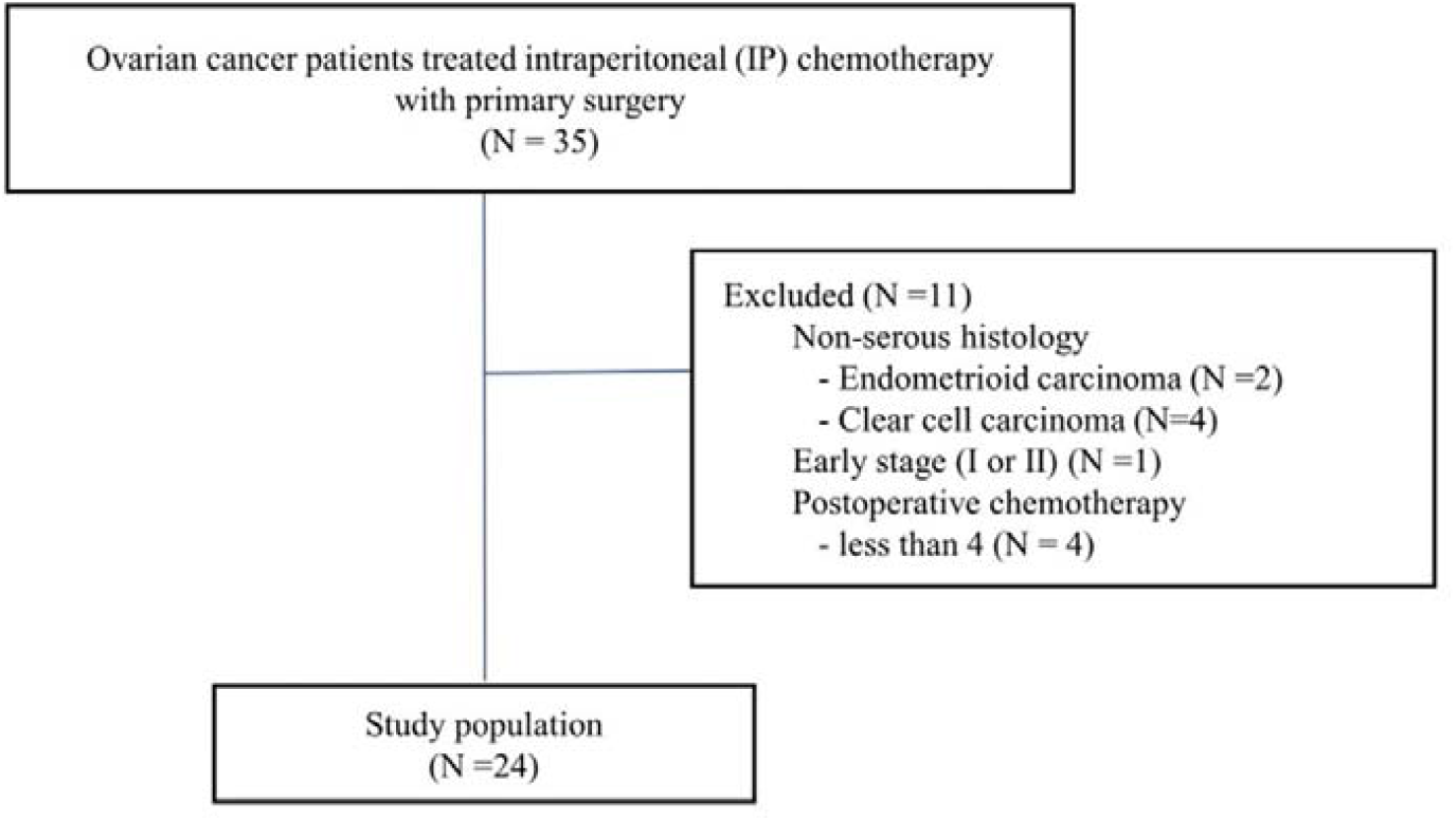
The flow of the study population.

### Selection of markers

Various markers, such as E-cadherin, vimentin, PD-L1, CD8 and FOXP3 are molecular markers that may play a crucial role as prognostic indicators and guide treatment decisions in ovarian cancer patients [7,8]. Recently, ovarian cancer therapies targeting tumor microenvironment (TME) is rapidly developing, targets mainly focusing on cancer-associated fibroblasts, tumor-associated macrophages, angiogenesis, and immune checkpoint blockade [3,9]. To date, no studies have investigated the association between the TME and survival outcomes in ovarian cancer patients treated with IP chemotherapy. Therefore, we aimed to analyze the relationship between factors such as CD8, FOXP3, PD-L1, E-cadherin, and Vimentin, and the response to IP chemotherapy.

### Immunohistochemistry

A board-certified pathologist (H.K) reviewed formalin-fixed paraffin-embedded tissue sections stained with hematoxylin and eosin to confirm the pathologic diagnosis and selected a representative paraffin block from each specimen for immunohistochemical analysis of surgically resected samples. Formalin-fixed, paraffin-embedded tissues were sectioned at a thickness of 4 μm and stained using an automated immunostainer (Ventana Medical Systems, Tucson, AZ, USA) according to the manufacturer’s protocol. The five antibodies and conditions used in this study – anti-CD8, FOXP3, PD-L1, E-cadherin,and vimentin – are listed in the Supplementary Table 1.

For TIL evaluation, CD8 and FOXP3-stained slides were scanned using a high-resolution digital slide scanner up to 400x magnification (3DHISTECH Pannoramic 250; 3DHISTECH Ltd., Budapest, Hungary) and counted automatically by a computerized image analysis system (QuantCenter 2.0; 3DHISTECH Ltd., Budapest, Hungary). The densities of cells expressing CD8 and FOXP3 were evaluated using NuclearQuant software that counted the positive cells throughout the entire tumor area but not in tissue outside the tumor border. The mean number of cells positive for each marker is expressed as density per mm^2^.

PD-L1 expression was defined if membranous and/or cytoplasmic staining was observed in tumor cells and tumor-associated immune cells. The combined positive score (CPS) was recorded based on the number of PD-L1-positive tumors and immune cells in relation to the total number of tumor cells. PD-L1 positivity was defined as a CPS > 1 [10]. E-cadherin and vimentin immunistaining were graded semiquantitatively based on the percentage of cells stained and the intensity of staining [11]. Briefly, the staining intensity was graded as weak (1+), moderate (2+), or strong (3+) and was multiplied by the percentage of positive cells. The total score was then classified as follows: 0-100 = grade 1, 101-200 = grade 2, and 201-300 = grade 3. Expression level was dichotomized using median value of scores for all 5 markers.

### Analysis

Descriptive statistics were employed to summarize the demographic data, which are presented as the median (range) or frequency (percentage). PFS was defined as the interval between the date of diagnosis and the date of first recurrence. Overall survival OS was defined as the interval between the date of diagnosis and the date of death. PFS and OS curves were estimated using the Kaplan-Meier method. The association between the levels of expression of the markers and survival outcomes was assessed using the log-rank test. A p-value of less than 0.05 was considered statistically significant. Statistical analyses were conducted using SPSS software, version 26.0 (IBM Corp., Armonk, NY).

## Results

### Clinicopathologic characteristics

The characteristics are shown in Table 1. The study included 24 patients with a median age of 44 years (ranging from 27 to 59). Most of the patients, 23 out of 24 (95.8%), were diagnosed at stage IIIC and all presented with high-grade serous histologic type. Residual disease larger than 1 cm was noted in 8 patients (33.3%). The predominant IP chemotherapy regimen was paclitaxel plus cisplatin (50.0%), and the median number of chemotherapy cycles was 6.8 (range 4 to 9). After IP chemotherapy, 20 patients (83.3%) experienced a recurrence. The median PFS for the cohort was 15.3 months (95% confidence interval (CI) range 0.4 to 148.3) and the OS was 63.3 months (95% CI, range 0.4 to 163+).

**Table 1.** Patient and clinical characteristics of the cohort (N=24)

| Characteristics | Patients<br>(N = 24) |
| --- | --- |
| Median age, years (range) | 44 (27-59) |
| FIGO stage, n (%) |  |
| IIIC | 23 (95.8%) |
| IVB | 1 (4.2%) |
| Histologic type, n (%) |  |
| Serous | 24 (100%) |
| Primary site, n (%) |  |
| Ovary | 22(91.7%) |
| Tube | 1 (4.2%) |
| Peritoneum | 1 (4.2%) |
| Residual disease, n (%) |  |
| No | 3 (12.5%) |
| <1cm | 13 (54.2%) |
| >1cm | 8 (33.3%) |
| IP Chemotherapy regimen, n (%) |  |
| Paclitaxel + cisplatin | 12 (50.0%) |
| Paclitaxel + carboplatin | 9 (37.5%) |
| Cisplatin | 1 (4.2%) |
| Carboplatin | 1 (4.2%) |
| Paclitaxel | 1 (4.2%) |
| Cycles of total chemotherapy,<br>median (range) | 6.8 (4-9) |
| Recurrence, n (%) |  |
| Yes | 20 (83.3%) |
| No | 4 (16.7%) |

IP, intraperitoneal; FIGO, International Federation of Gynecology and Obstetrics.

### Association of markers with survival

None of the five markers—CD8, FOXP3, PD-L1, E-cadherin, and Vimentin—demonstrated a statistically significant association with either PFS or OS. The respective p-values for each marker were as follows: CD8 (p=0.63), FOXP3 (p=0.70), PD-L1 (p=0.98), E-cadherin (p=0.37), and Vimentin (p=0.92). The p-values for each marker in relation to overall survival (OS) were as follows: CD8 (p=0.20), FOXP3 (p=0.57), PD-L1 (p=0.96), E-cadherin (p=0.92), and Vimentin (p=0.10).

Given the marginal associations of CD8 (p=0.20) and vimentin (p=0.10) with OS, patients were stratified into two groups: those with high expression of either CD8 or vimentin (n=18) and those with low expression of both markers (n=6). Patients with high expression of CD8 or vimentin demonstrated a numerically longer PFS (median: 19.7 months vs. 13 months, p=0.073) and a significantly improved OS (median: 94.5 months vs. 25.4 months, p=0.008) compared to those with low expression of both markers, as shown in Fig. 2.

**Figure 2.**
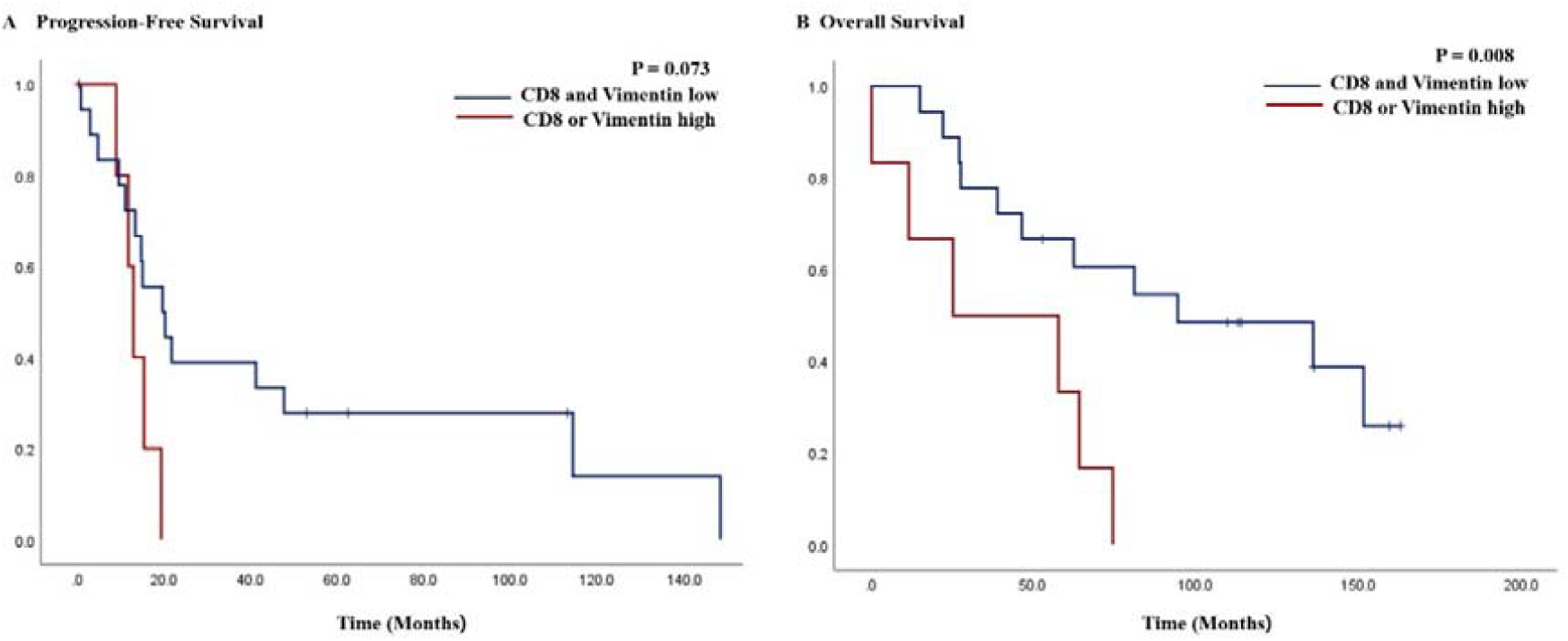
Kaplan-Meier curves of progression-free survival (A) and overall survival (B) in patients with high expression of CD8 or vimentin vs. low expression in both CD8 and vimentin.

## Discussion

The rationale for IP chemotherapy is based on the fact that ovarian cancer commonly spreads within the peritoneal cavity., even after extensive cytoreductive surgery [8]. The Gynecologic Oncology Group (GOG) has shown through three large randomized, phase III clinical trials (GOG 104, 114, and 172) that IP chemotherapy can offer superior PFS and OS compared to IV chemotherapy [9,12,13]. However, there were no clear studies of molecular markers expression affecting survival in ovarian cancer patients treated with IP chemotherapy. Our study demonstrated that higher levels of CD8 and vimentin expression were associated with significantly improved survival outcomes in ovarian cancer patients receiving IP chemotherapy. This finding suggests that the combined analysis of CD8 and Vimentin may provide deeper insights into the effectiveness of IP chemotherapy for ovarian cancer. To the best of our knowledge, this study is the first report identifying molecular markers associated with response to IP chemotherapy in ovarian cancer.

CD8, a marker for cytotoxic T lymphocytes, has been identified as a positive prognostic factor in various cancers, including ovarian cancer. Previous studies showed that, Vimentin, a marker of epithelial-mesenchymal transition, is associated with increased tumor aggressiveness and metastatic potential [14-16]. However, there are currently no studies on its association with IP chemotherapy.

Tumor mRNA expression was used to identify genes that confer survival benefits following IP chemotherapy [17,18]. The recently published study showed that hyperthermic intraperitoneal chemotherapy (HIPEC) plays a role in modulating the TME changes. These studies included the biomarker analysis that employed whole-exome and whole-transcriptome sequencing changes after HIPEC [18,19]. Despite of the extensive gene expression analysis, both studies did not confirm tumor protein expression experimentally by immunohistochemistry of primary tumor specimens, which is a major difference from our study. Changes in protein expression can be observed relatively faster with immunohistochemical analysis. This approach may facilitate the optimization of treatment strategies, enabling the timely administration of appropriate therapies and identifying the subset of patients most likely to benefit from IP chemotherapy.

The limitations of this study include its retrospective design and the small sample size, which may impact the robustness of the findings. Additionally, the binary classification of marker expression levels based on median values may oversimplify the complex biological roles of these markers in ovarian cancer. Notably, unlike OS, none of the five markers (CD8, FOXP3, PD-L1, E-cadherin, and Vimentin) demonstrated a significant association with PFS. This discrepancy suggests that these biomarkers may have a more complex role in disease progression. Further investigation in larger cohorts is necessary to validate these findings and elucidate the complex interplay between these markers and disease dynamics. Future studies with larger sample sizes are warranted to confirm these results and explore the underlying biological mechanisms. We demonstrated that the expression of CD8 and vimentin is associated with OS in ovarian cancer patients who received IP chemotherapy. To our knowledge, this is the first study to experimentally validate the response to IP chemotherapy based on specific protein expression analyzed through immunohistochemistry of primary tumor specimens. These findings suggest a potential approach to identifying patients most likely to benefit from IP chemotherapy, paving the way for more targeted and effective therapeutic strategies. Further research is required to elucidate biomarkers predictive of favorable outcomes with IP chemotherapy.

## Data Availability

All data produced in the present study are available upon reasonable request to the authors

## Acknowledgments

None

---

**Supplementary Table 1.**
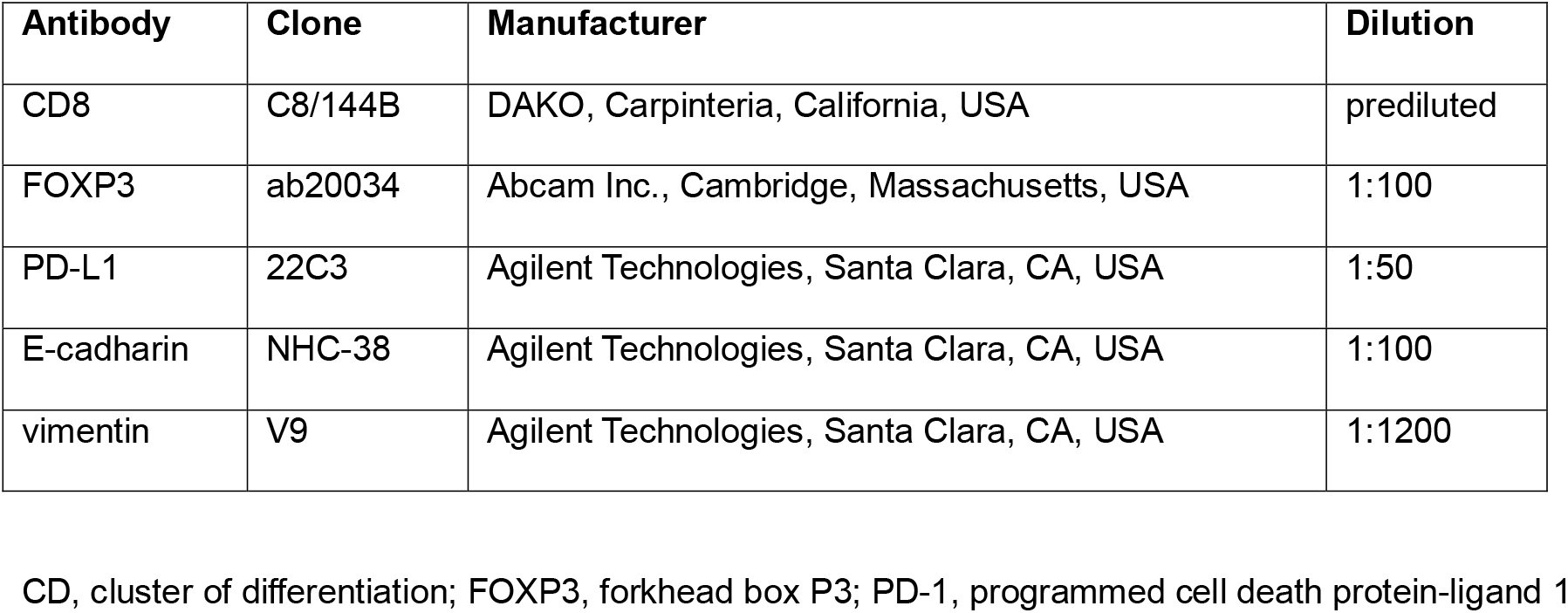
Antibodies and conditions used in the study. CD, cluster of differentiation; FOXP3, forkhead box P3; PD-1, programmed cell death protein-ligand 1

## References

1. Lee JY, Kim EY, Jung KW, Shin A, Chan KK, Aoki D, et al. Trends in gynecologic cancer mortality in East Asian regions. J Gynecol Oncol 2014;25:174–82.

2. Colombo N, Lorusso D, Scollo P. Impact of Recurrence of Ovarian Cancer on Quality of Life and Outlook for the Future. Int J Gynecol Cancer 2017;27:1134–40.

3. Lee YJ, Woo HY, Kim YN, Park J, Nam EJ, Kim SW, et al. Dynamics of the Tumor Immune Microenvironment during Neoadjuvant Chemotherapy of High-Grade Serous Ovarian Cancer. Cancers (Basel) 2022;14

4. Walker JL, Brady MF, Wenzel L, Fleming GF, Huang HQ, DiSilvestro PA, et al. Randomized Trial of Intravenous Versus Intraperitoneal Chemotherapy Plus Bevacizumab in Advanced Ovarian Carcinoma: An NRG Oncology/Gynecologic Oncology Group Study. J Clin Oncol 2019;37:1380–90.

5. Shi T, Jiang R, Pu H, Yang H, Tu D, Dai Z, et al. Survival benefits of dose-dense early postoperative intraperitoneal chemotherapy in front-line therapy for advanced ovarian cancer: a randomised controlled study. Br J Cancer 2019;121:425–8.

6. Jaaback K, Johnson N, Lawrie TA. Intraperitoneal chemotherapy for the initial management of primary epithelial ovarian cancer. Cochrane Database Syst Rev 2011:Cd005340.

7. Lee JY, Lee YY, Park JY, Shim SH, Kim SI, Kong TW, et al. Major clinical research advances in gynecologic cancer in 2022: highlight on late-line PARP inhibitor withdrawal in ovarian cancer, the impact of ARIEL-4, and SOLO-3. J Gynecol Oncol 2023;34:e51.

8. Cannistra SA. Cancer of the ovary. New England Journal of Medicine 2004;351:2519–29.

9. Singhal P, Lele S. Intraperitoneal chemotherapy for ovarian cancer: where are we now? Journal of the National Comprehensive Cancer Network 2006;4:941–6.

10. Kim Y, Aiob A, Kim H, Suh DH, Kim K, Kim YB, et al. Clinical Implication of PD-L1 Expression in Patients with Endometrial Cancer. Biomedicines 2023;11

11. Park E, Park SY, Sun PL, Jin Y, Kim JE, Jheon S, et al. Prognostic significance of stem cell-related marker expression and its correlation with histologic subtypes in lung adenocarcinoma. Oncotarget 2016;7:42502–12.

12. Markman M, Bundy BN, Alberts DS, Fowler JM, Clark-Pearson DL, Carson LF, et al. Phase III trial of standard-dose intravenous cisplatin plus paclitaxel versus moderately high-dose carboplatin followed by intravenous paclitaxel and intraperitoneal cisplatin in small-volume stage III ovarian carcinoma: an intergroup study of the Gynecologic Oncology Group, Southwestern Oncology Group, and Eastern Cooperative Oncology Group. Journal of clinical oncology 2001;19:1001–7.

13. Armstrong DK, Bundy B, Wenzel L, Huang HQ, Baergen R, Lele S, et al. Intraperitoneal cisplatin and paclitaxel in ovarian cancer. New England Journal of Medicine 2006;354:34–43.

14. Bachmayr-Heyda A, Aust S, Heinze G, Polterauer S, Grimm C, Braicu EI, et al. Prognostic impact of tumor infiltrating CD8+ T cells in association with cell proliferation in ovarian cancer patients-a study of the OVCAD consortium. BMC cancer 2013;13:1–8.

15. Khashaba M, Fawzy M, Abdel-Aziz A, Eladawei G, Nagib R. Subtyping of high grade serous ovarian carcinoma: histopathological and immunohistochemical approach. Journal of the Egyptian National Cancer Institute 2022;34:6.

16. Szubert S, Koper K, Dutsch-Wicherek MM, Jozwicki W. High tumor cell vimentin expression indicates prolonged survival in patients with ovarian malignant tumors. Ginekologia Polska 2019;90:11–9.

17. Seagle B-LL, Eng KH, Yeh JY, Dandapani M, Schiller E, Samuelson R, et al. Discovery of candidate tumor biomarkers for treatment with intraperitoneal chemotherapy for ovarian cancer. Scientific reports 2016;6:21591.

18. Dellinger TH, Han ES, Raoof M, Lee B, Wu X, Cho H, et al. Hyperthermic Intraperitoneal Chemotherapy–Induced Molecular Changes in Humans Validate Preclinical Data in Ovarian Cancer. JCO Precision Oncology 2022;6:e2100239.

19. Eoh KJ, Lee J-Y, Nam EJ, Kim S, Kim YT, Kim SW. Long-term survival analysis of intraperitoneal versus intravenous chemotherapy for primary ovarian cancer and comparison between carboplatin-and cisplatin-based intraperitoneal chemotherapy. Journal of Korean medical science 2017;32:2021.

